# The Covid-19 containment effects of public health measures - A spatial difference-in-differences approach

**DOI:** 10.1101/2020.12.15.20248173

**Authors:** Reinhold Kosfeld, Timo Mitze, Johannes Rode, Klaus Wälde

## Abstract

Since mid-March 2020 the Federal and state governments in Germany agreed on comprehensive public health measures to curb the spread of SARS-CoV-2 infections leading to the Covid-19 disease. We study the containment effects of these policy interventions on the progression of the pandemic in the first containment phase in spring 2020 before the easing of restrictions may become effective by the end of April. To exploit both the temporal and spatial dimension in the dissemination of the virus, we conduct a spatial panel data analysis for German NUTS-3 regions. Specifically, we employ a spatial difference-in-differences approach to identify the effects of six compound sets of public health measures. We find that contact restrictions and closure of schools substantially contributed to flattening the infection curve. Additionally, a strong treatment effect of mandatory wearing of face masks is established for the few treated regions during this containment phase. No incremental effect is evidenced for closure of establishments, such as museums, theaters, cinemas and parks, and the shutdown of shopping malls and other non-essential retail stores. These findings prove to be robust to changes in model specification. By contrast, the dampening effect of restaurant closure is sensitive to model variation.

## 1 Introduction

The disruptions caused by the Covid-19 pandemic might turn out to be greater than any other event since the Second World War. Governments and authorities around the world therefore have taken drastic measures since early 2020 to slow down the pandemic. Measures include the closing of schools and non-essential sales shops, mandatory face masks, travel and contact restrictions and even contact bans or curfews. It is no surprise that these measures encounter partially fierce protests by some politicians and parts of the public. It is high time to understand how effective the various measures were in reducing the spread of Covid-19 to be prepared for future epidemic and pandemic developments. This is the purpose of this paper.

Evaluating the effectiveness of public health measures is not trivial. When we take the example of Germany, there was initially a strong political consensus that policy measures should be identical across federal states. In fact, apart from some exceptions, first measures decided upon on Friday March 13 were introduced across federal states in a relatively uniform matter. Yet, given the federal structure of Germany, some variation still exists. The regulations were usually enacted at the level of federal states. For each of the six compound sets of public health measures we are analyzing, we find differences in the timing of implementation of up to one week. Additionally, in a few local administrative regions within the states, specific regulations came into force on different dates.

We exploit this variation to identify the effect of these public health interventions on the spread of Covid-19. We start the period of investigation at the beginning of the last week in February, where positive SARS-CoV-2 infection counts are registered in the vast majority of regions. Depending on the specific policy intervention in focus, the pre-treatment period accordingly lasts between three and four weeks. As there is evidence that immediate mitigating effects are stronger than easing impacts (Deb et al. 2020), the evaluation period terminates at the end of the penultimate week of April before first easing effects should become visible in the data.

We employ a spatial difference-in-differences (DiD) approach (Delgado and Florax 2015; Chagas, Azoni and Almeida 2016) to identify the effects of public health measures. To exploit both the temporal and spatial dimension in the dissemination of SARS-CoV-2/Covid-19 (thereafter: Covid-19), we conduct a spatial panel data analysis for 401 German NUTS-3 regions. A spatialisation of the usual DiD estimator is necessary to avoid biased and inconsistent treatment effects in the presence of substantial spatial spillovers (Kolak and Anselin 2020). Pure spatial regressions fail to establish causal effects. Being relatively new, up to now – to the best of our knowledge – no other Covid-19 related studies took advantage of the benefits of the spatial DiD approach.

We use official data on registered daily Covid-19 case counts for the sample period from 24 February to 24 April 2020. Taking account for a pre-treatment period of at least three weeks, this time frame covers the containment phase that ends when first easing measures can be expected to become effective. In order to account for regional spillovers in the spread of infections and local treatment effects, data analysis is performed in the framework of the spatial Durbin model (SDM). For the purpose of comparison, we disregard both types of spatial effects in the baseline DiD model. To check the robustness of identifying causal effects, we also make use of alternative spatial panel data models. Given the temporal and regional variation in their implementation, we analyze the effectiveness of six compound sets of containment measures.

The study reveals considerable differences in the potency of implemented public health interventions to flatten the pandemic curve in Germany. Contact restrictions as an essential means of social distancing are shown to be most effective in dampening the growth of regional infection rates. On average, their highly significant incremental impact amounts to nearly 14 percentage points of the cumulative incidence growth rate. The estimated treatment effect of 13 ½ percentage points of mandatory wearing of face masks in public transportation and sales shops is only based on a few regions being treated during the containment phase. In consequence, the impact of the face mask duty can be estimated with less precision in our sample settings than that of contact restrictions (see Mitze et al. 2020 for an alternative study design focusing specifically on the Covid-19 effects of face masks). A highly significant dampening effect of 5 ½ percentage point is also found for the closure of schools and daycare facilities. No significant incremental effects are identified for closure of establishments (cinemas, theaters, museums etc.) and the shutdown of non-essential retail stores in shopping malls and other locations. These findings are robust with respect to various changes in model specification. By contrast, the negative significant impact of restaurant closure on the progress of the first Covid-19 pandemic wave in Germany turns out to be sensitive to model variation.

The remainder of the paper is structured as follows. With a special focus on Covid-19 studies, section 2 reviews the literature on impact of non-pharmaceutical interventions to curb infectious diseases. In section 3, public health measures for Covid-19 prevention are outlined and data are described. The spatial differences-in-differences (DiD) approach is introduced in section 4. In section 5, empirical findings on the effectiveness of the measures are presented and discussed. Robustness checks are conducted in section 6. Section 7 concludes this study.

## 2 Literature Review

This section reviews the available prior evidence on the effectiveness of non-pharmaceutical public health interventions to suppress the spread of Covid-19 in Germany. To approach this issue in a systematic manner (against the background of a considerable number of research contributions on Covid-19), we mainly confine our literature review to studies that incorporate an explicit regional dimension and focus on those papers that have already undergone a peer review process. Complementary studies at the aggregate national level, which deal with the question if governmental restrictions to public and private life have managed to “flatten” Germany’s infection curve during the first wave of the pandemic in spring 2020, can, for instance, be found in Dehning et al. (2020), Hartl et al. (2020), Donsimoni et al. (2020a, 2020b), Glogowsky et al. (2020), and Wieland (2020a). These studies mainly conclude that policy interventions, particularly the full lockdown (including contact bans) implemented on March 23 helped to significantly slow down the spread of Covid-19 – although some studies, such as Wieland (2020a), find that a decline in the infection dynamics may already have taken place prior to this date, i.e. in early March when large events have been banned.

A potential limitation of aggregate data studies that track the development of the German-wide infection curve is that these studies typically use an indirect identification approach, based on tests for structural breaks in the aggregate data over time which is then associated with a certain policy event or group policy interventions into few or even a singular policy “lockdown” shock(s). This limits the interpretation in terms of the underlying transmission mechanisms and prevents the studies from differentiating between the specific effects of alternative policy measures implemented in a staggered way across space and over time. One notable exceptions is the international comparison study of Flaxman et al. (2020) which accounts for the latter by estimating disaggregate effects for different types of non-pharmaceutical interventions (including full lockdown, ban on public event, school closure, self-isolation and social distancing) on Covid-19 reproduction and death rates in European countries (including Germany). Similar to the above-mentioned single country studies for Germany, this cross-country approach finds that large-scale lockdowns of populations have an identifiable large effect on disease transmission, i.e. reduce Covid-19 infection rates by approx. 81%. However, with regard to the attempt of disentangling the effects of different policy measures, the authors conclude that the close spacing of interventions in time does not allow to properly identify the individual effects of other interventions than large-scale lockdowns.

From an empirical identification perspective, the estimation results reported in Flaxman et al. (2020) provide an argument for using regional rather than national data when assessing the effectiveness of non-pharmaceutical interventions in the context of Germany. Given the federal organization of German health policy with state-level governments in the 16 federal states (Bundesländer) being in charge for the design and implementation of public health measures, this specific institutional setup allows the researcher to exploit space-time discontinuities in the implementation of non-pharmaceutical interventions, which may overcome the above-mentioned identification problems reported in Flaxman et al. (2020). Besides, the use of disaggregate data below the national level can account for spatial heterogeneities in regional infection dynamics and consider spatial transmission processes across regions as an important mechanism of disease spread when assessing the effectiveness of public interventions on Covid-19.

There are some earlier papers that have already exploited the regional dimension when modelling Covid-19 infections in Germany. However, only very few have placed a distinct focus on studying the effectiveness of policy interventions by systematically exploiting space-time discontinuities therein. The studies that are most closely related to our approach taken here are Berlemann and Haustein (2020), Wieland (2020b) and Kergassner et al. (2020). All three studies encompass in the spatio-temporal dynamics of the Covid-19 epidemic in Germany - both in terms of data and estimation. Berlemann and Haustein (2020) use data at the district level (NUTS3 regions) to estimate an endemic-epidemic model for regional counts of newly infected persons. Their empirical specification chiefly builds on an autoregressive process of disease dynamics augmented by spatial lag terms (comprising the epidemic part of the model) together with a deterministic time trend (endemic part) and random regional effects. The authors use this model to evaluate the effectiveness of three waves of containment measures, namely 1) ban of mass events, 2) closure of educational institutions, leisure facilities and retail facilities together with and international traveling restrictions, 3) social distancing (contact restrictions) on the spread of Covid-19 within and across German districts.

The estimation results reported by Berlemann and Haustein indicate that only the first wave of containment measures clearly contributed to flatten the curve of new infections; however, the authors acknowledge that interpretations need to be done only carefully as in their empirical setup the estimated effect of the adopted measures is contingent on which measures were adopted before. Although Berlemann and Haustein do not explicitly consider space-time discontinuities in the implementation of policy interventions across federal states, their analysis of regional responses to a common intervention shock further indicates that a one size-fits-all policy is not optimal from a regional perspective, i.e. implemented policy measures helped to slowdown the infection dynamics in those federal states that were most severely affected by Covid-19 but did note change infection trajectories in other states. Similar to Berlemann and Haustein, the results reported in Wieland (2020b) provide mixed evidence for a regional trend change in infections around the timing of non-pharmaceutical inter¬ventions. As the author points out, in nearly two thirds of German counties the flattening of the infection curve was found to occur before the full lockdown came into force on March 23 and one in eight counties even experienced a decline of infections even before the closures of schools, child day care facilities and retail stores.

Kergassner et al. (2020) model the regional infection dynamics in Germany considering the three waves of containment measures as studied by Berlemann and Haustein and, in addition, account for federal-state-specific (staggered) starting dates of these measures. The results from a coupled SIQRD epidemic and mobility network model show that infection rates during the time period considered show a significant degree of heterogeneity across federal states and, accounting for this spatial heterogeneity, significantly improves model predictions at the federal state and district level. Kergassner et al. use the spatially resolved model to infer infection effects stemming from selected seeds such as 1) Heinsberg in Germany during the Carnival season and 2) returning travelers from Ischgl in Austria. Based on their empirical findings the authors conclude that refraining from traveling and large events are two key interventions that can effectively attenuate the spreading of Covid-19 at the subnational level. Felbermayr et al. (2020) similarly report that particular types of travel, i.e. holiday returnees from skiing vacations, proxied by the road distance to Ischgl, constitute an important predictor of infection cases across German districts. When assessing the superspreader effects emanating from the Ischgl Covid-19 outbreak over time, Felbermayr et al. find that restrictions in mobility after March 23 have helped contain the virus imported from Ischgl in those counties where it first arrived.

While the above approaches broadly account for the role of spatial transmission, no detailed account is given on the specific transmission mechanisms across space. This is done in Bluhm and Pinkovskiy (2020) and Mitze and Kosfeld (2021), who investigate the role of commuting flows across regions as a determinant for the regional evolution of Covid-19 cases. Blum and Pinkovskiy particularly focus on analysing discontinuities in Covid-19 cases along the West-East internal German border. Starting from the hypothesis that the observed macro-regional heterogeneities between West and East Germany may be driven by different vaccination policies during the Cold War era (especially linked to the BCG vaccine against tuberculosis), in the course of their empirical investigation the authors do not find empirical support for this BCG hypothesis but show that commuting patterns and regional demographic structures are significant factors explaining regional differences in Covid-19 cases and deaths per capita.

A significant propagation effect of commuting to work is also found in Mitze and Kosfeld (2021) when employing a spatial econometric model to analyze the spatio-temporal dynamics of Covid-19 cases at the district level (NUTS-3 regions). The authors further find that this commuting channel of disease transmission breaks down after the imposition of professional and social contact constraints during the German lockdown period after March 23. Another route is taken in Fritz and Kauermann (2020), who study the role of mobility and social connectivity derived from Facebook activities on the spatio-temporal dynamics of Covid-19 cases at the district level in Germany. Fritz and Kauermann find evidence for a significant correlation between reduced social activity and lower infection rates and that the extent of social distancing has an overall negative effect on the incidence of infections.

Finally, Huber and Langen (2020) study the effect of mitigating public health measures on cumulative Covid-related hospitalization and death rates in Germany and Switzerland. For effect identification, the study exploits the fact that the epidemic was more advanced in some regions than in others when certain lockdown measures came into force. This difference is used to define alternative treatment groups based on the days between the district-specific start of the epidemic and the lockdown measured through non-essential retail shop closure between March 17 and 20, which are grouped into an early, intermediate and late intervention group. The results indicate that regions in the late intervention group, i.e. those regions that have experienced a longer temporal lag between the start of the local infection and the German-wide lockdown, indeed come with higher death rates. Huber and Langen attribute the estimated effect to federally imposed contact restriction but find no additional effect for local curfews introduced in some federal states.

One particular strength of the study of Huber and Langen is the combination of multiple data datasets and estimation routines, which allows the authors to assess the robustness of the estimated effects. For the sample of German regions, Huber and Langen do so by applying both simple OLS and doubly robust estimation to account for the endogeneity of the treatment. The authors also control for district-specific characteristics and state-specific policy measures entering into force prior to the general lockdown. However, a limitation is that regressions are only run in a cross-sectional setup and that spatial effects are not considered. We will explicitly consider both of these limitations in the estimation of spatial difference-in-differences models to comprehensively assess the effectiveness of non-pharmaceutical interventions in Germany.

## 3 Containment Measures and Data

There are 16 federal states (NUTS-1 regions) and 401 NUTS-3 districts in Germany. In Table 5 of Appendix A, we quote the number NUTS-3 regions (districts) in each federal state.

### 3.1 Containment Measures

We make use of six compound categories of containment measures. In Table 5 of Appendix A, we show the dates of their implementation. In most cases, the containment measures were put into force at the federal state level. If this is the case, the measure is binding for all NUTS-3 regions within the corresponding federal state. We did our best to check for differing implementation dates in specific NUTS-3 regions. If we found differences, we have accounted for them and state them in the following.

#### School closure

This containment measure also refers to other educational institutions (e.g. universities) and daycare facilities (kindergartens). Again, we exploit that federal states did not implement the containment measure at the same time. Bavaria, Bremen, Hamburg, Mecklenburg-West Pomerania, Rhineland-Palatinate, Saxony-Anhalt and Schleswig-Holstein closed schools on March 16. The treatment group thus includes 296 districts for this specific date. Baden-Wurttemberg, Berlin and Thuringia followed on March 17, which extends the treatment group to 364 districts from this date onwards. Brandenburg, Saarland and Saxony were last and closed schools on March 18. In some districts, schools were already closed on March 13, which means that these districts receive treatment early on.

#### Establishment closure

This category of containment measures refers to the closure of public and private establishments including museums, theaters, cinemas, swimming pools, fitness studios, zoological gardens, public parks and playgrounds. We use differences in the date of implementation in federal states to ascertain treatment effects of this compound measure. Berlin and Schleswig Holstein were the first to close establishments on March 15. For a limited period of time, our treatment group thus only consists of 16 districts only. Hamburg and North Rhine-Westphalia followed on March 16 extending the treatment group to 70 districts from this day onwards. Brandenburg, Saarland and Saxony were last and closed establishments on March 19. The remaining states implemented the containment measure on March 17 or 18.

#### Shopping mall and non-essential retail store closure

According to the regulations on business operations, shopping malls and further non-essential retail stores (excluding pharmacies, building centers, drug and food stores) had to be closed. Business shutdown also applied to household-related service activities like hairdressers, cleaners and launderettes. Shopping malls and other stores were closed first in Hamburg and Lower Saxony on March 17. For this date, 46 districts are included in the treatment group. While 11 other federal states followed on March 18, Saxony closed non-essential retail stores on March 19, Thuringia on March 20 and Berlin on March 23.

#### Restaurant closure

Restaurants, bars and further leisure facilities (e.g. bars and clubs) were closed in German federal states from March 18 onwards starting with Schleswig-Holstein. For two days our treatment group thus consists of 15 districts. Lower Saxony (45 districts) and Thuringia (23 districts) followed on March 20. Nine of the remaining states implemented the containment measures on March 21. While Berlin followed on March 22, Brandenburg and North Rhine-Westphalia waited until March 23. Saxony-Anhalt was last and closed restaurants on March 25.

#### Contact restrictions

We estimate treatment effects from contact restrictions by exploiting the early implementation of this containment measure in two states. While Bavaria and the Saarland put the measure into force on March 21, the Federation-State Agreement for all German states was announced on the evening of March 22. Thus, for a short period of time, the 102 districts of Bavaria and Saarland constitute the treatment group, whereas the 300 districts of all other German states serve as comparison regions receiving treatment only later on. While in most German states the regulation on contact restrictions came into force in accordance to the Federation-Länder Agreement (Bund-Länder-Vereinbarung) on March 23, it was officially enforced on March 24 in Rhineland-Palatinate and on March 25 in Thuringia and Saxony-Anhalt.

#### Face mask duty

Saxony was first federal state to make face masks mandatory (in public transport and sales shops) on April 20. Saxony-Anhalt followed with its 14 districts two days later and Thuringia 4 days later with another 23 districts joining the treatment group in a staggered manner. The remaining states (but Schleswig-Holstein) made face masks mandatory on April 27. Schleswig-Holstein implemented the obligation to wear face masks on April 29. Face masks became mandatory early only in some NUTS-3 districts: Jena (Thuringia) was first on April 6, Nordhausen (Thuringia) followed on April 14, Landkreis Rottweil (Baden Wurttemberg) on April 17, and Main-Kinzig-Kreis (Hesse) and Wolfsburg (Lower Saxony) on April 20. In consequence, our treatment group consists of only one district between April 6 and April 13. For the following three days, it includes two districts, and for another three days, three districts.

While the above containment measures were designed to contain the spread of Covid-19 on a continuous daily basis from their date of implementation onwards, potential effects of other measures such as the ban of major sporting events from mid-March onwards mainly applied to weekends. Other large events like festivals that were prohibited by state governments usually as well take place on weekends. As the ban of major events were effective around the same time as the school closures, a distinct effect of mass events is difficult to identify (cf. Flaxman et al. 2020; Von Bismarck-Osten, Borusyak and Schönberg 2020).^2^ The identification problem is aggravated by the fact that daycare facilities and schools were closed earlier in certain NUTS-3 regions. Because closing of educational institutions affects a considerably larger share of population as a ban of major events, we put the closure of daycare facilities and schools in the foreground. As a consequence of this latent identification problem, the estimated impact of school closures might partly catch effects from bans on mass events and therefore may overestimate the “true” effect of school closure. We carefully address this aspect in the presentation of the empirical results.

As shown above, the implementation of the above categories of containment measures took place over a time interval between March 15 to April 29 exhibiting a certain degree of spatio-temporal variation in treatment receipt. For a convincing empirical analysis, i.e. to capture trend developments prior the the start of these measures, we need a sufficiently long pre-treatment period. That is why, our period of analysis starts three weeks before the first implementation of a containment measure on March 15. Because of expected asymmetries (Deb et al. 2020), we have to assure that there is no relevant overlap between the implementation and easing of containment measures during our investigation period. The first easing measure has been introduced on April 20. On that day, Brandenburg re-opened establishments and all federal states (except Bavaria) allowed non-essential retail stores to re-open (under certain restrictions). Mitze et al. (2020) report a median delay of 10.5 days (and 5.7 days for the 10*th* percentile) until a change in a specific containment measure can be expected to be visible in the reported Covid-19 cases in Germany. While using a longer time dimension of our sample period would be generally preferable, in order to avoid an overlap with the gradual phasing-in of easing measures, we are cautious regarding overlapping effects and end our analysis on April 24.

### 3.2 Data

We employ the official German regional statistics on reported Covid-19 cases from the Robert Koch Institute (RKI, 2020a). We build a balanced panel for 401 NUTS-3 districts and 61 days spanning the period from February 24 to April 24, 2020 (24,461 observations). Figure 1 illustrates the spatial distribution of cumulative reported Covid-19 cases at the beginning, in the mid and at the end of our period of analysis. On February 24, there were only 13 NUTS-3 regions with one or more reported cases. In the following month, we observe a large increase. On March 24, ten districts had more than 400 cumulative cases: Hamburg had most and reported 1617, Munich followed with 1571 and Berlin was third with 1501. At the same time, 91 districts had less than 25 cases. At the end of the investigation period, we see only in nine NUTS-3 regions less than 25 cumulative reported cases. Most of the districts with a low number lie in the north or east of Germany. Again, the largest cities reported the highest number of cumulative cases: Berlin reported 5600, Munich 5464 and Hamburg 4705 cases.

**Figure 1:**
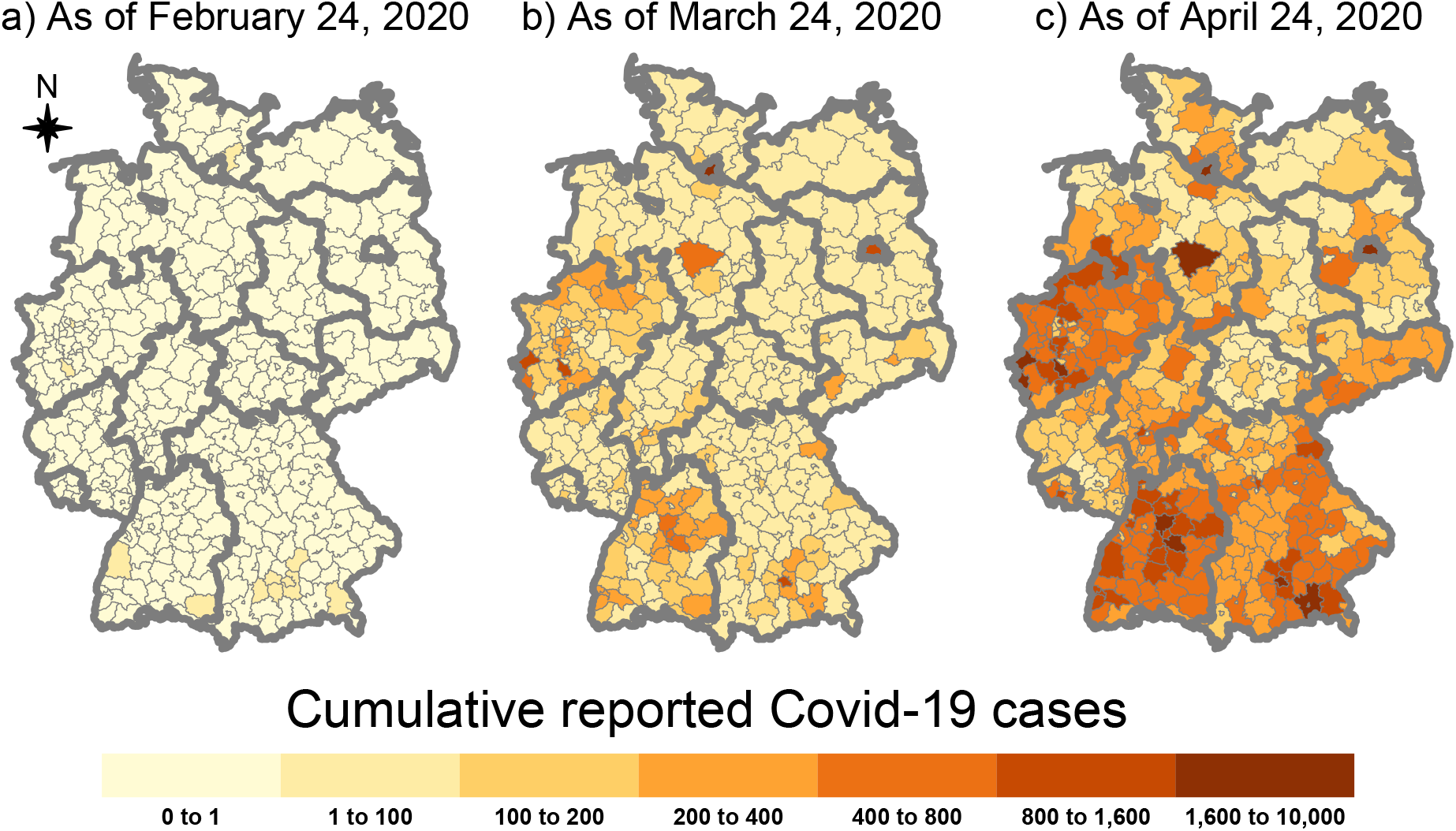
Cumulative reported COVID-19 Cases at the beginning and the end of our period of analysis at the NUTS-3 level. *Notes:* The data comes from RKI (2020a). Our unit of analysis are 401 NUTS-3 regions. Bold greylines indicate the borders of the 16 federal states.

We wish to establish impacts of the containment measures on the growth of the pandemic curve. To account for varying sizes of NUTS 3 regions, we normalize the number of daily Covid-19 cases by population of each district. The normalized cumulative cases are used to calculate the logarithmic cumulative incidence rates for district *r* at day *t*:

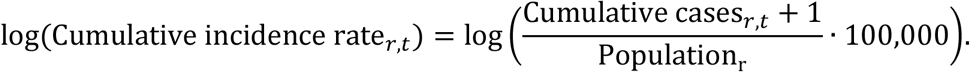

The cumulative incidence growth rate is formed as the difference of temporally consecutive values of the logarithms of cumulative Covid-19 incidence rates:

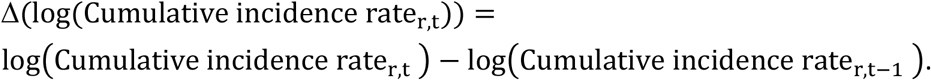

Summary statistics of Covid-19 variables and containment measures (binary dummies) are reported in **Fehler! Verweisquelle konnte nicht gefunden werden**. German authorities (RKI, 2020a) measure the number of reported infections by day of reporting and by day of appearance of first symptoms. Because around one third of the observations lack accurate information on the day of first symptoms, we use the day of reporting in our study. In Appendix B, we discuss the advantages and disadvantages of this approach in detail. The containment measures provide information on their length of implementation during the period of investigation.

## 4 Econometric Modelling

In assessing the effectiveness of policy measures, their impacts are often established from the differences between the treatment and control group after and before an action (Angrist and Pischke 2009, p. 169-174). When measures are adopted at different points in time, policy impacts can not only be exploited from variations between different entities at the same time but as well as from temporal changes of the variable of interest due to group changes of entities (Goodman-Bacon 2018). Public health measures to curb the spread of Covid-19 have been implemented in a staggered manner over time. Difference-in-differences (DiD) approaches can be founded on a panel model approach in order to analyze incremental treatment effects of static and dynamic type (Wooldridge 2010, p. 968-975).

Standard DiD estimators presuppose that a treatment of a unit does not affect the outcome of another unit. This observation rule is implied by the stable unit treatment value (SUTVA) assumption that excludes any kind of spillover effects from treated units (Angrist, Imbens and Rubin 1996; Wooldridge 2010, 509). The SUTVA assumption is clearly violated when people interact with one another across regional borders. As Covid-19 has spread across countries and regions, effective actions in one region are expected to exert dampening effects in surrounding areas. As infection spillovers are not restricted to local surroundings, we have equally to account for global spatial spillovers to identify causal effects of the public health measures. This demands a spatialization of the DiD approach in establishing treatment effects from panel data models with interacting regions.

In view of the different sizes of regions, we make use of incidence rates instead of the registered numbers of infections. Specifically, we utilize the logarithmic differences of cumulative infections rates, Δcir_rt_, as the dependent variable of the panel data models. The subscript r∈ {r=1,2,…,n} is the region index and t∈{1,2,…,T} the day number. In order to establish the causal effects of K public health measures, we define K intervention variables 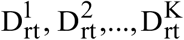 that take the value 1 if action k is enforced in region r at day t. Otherwise, the variable 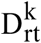 takes the value 0. In the case of regional interaction, particularly due to commuter flows within travel-to-work regions, neighbouring districts will as well be affected by the policy actions. Conversely, infection growth in a region will be impacted by the introduction of effective containment measures in its local surroundings. In such a case, a spatial lag 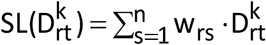 with w_rs_ as elements of the nxn spatial weights matrix **W** = [w_rs_]_nxn_ has to be included in the panel model.

The spatial weights matrix provides an operationalisation of connectivity of regions in space. The can be accomplished by using contiguity-based or distance-based weights (Anselin and Bera 1998, 243; Arbia 2006, 37-38). For the application of the contiguity approach^3^, we define the elements of a binary weights matrix **W**^*^ by

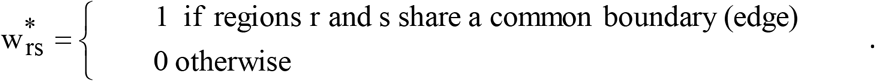

The spatial weights 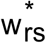 are 0 for r=s and non-adjacent regions. With row-standardised weights, 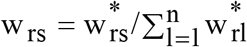, the spatial lag 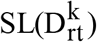 gives the share of treated areas with regard to policy action k in the surrounding of region r at day t.

As is well known from the usual DiD approach, the parallel trend assumption must be met for a valid identification of treatment effects. This assumption presupposes homogeneity of infection dynamics across regions in the pre-treatment period. In the presence of heterogenous trajectories^4^, we accommodate trend differentials by including region-specific trends β_r_ · t in the panel model (Angrist and Pischke 2015; Jaeger, Joyce and Kaestner 2018; Deb et al. 2020). Controlling for region fixed effects α_i_, time fixed effects δ_t_, and daily seasonalities (day_j_)^5^, the panel data model underlying the localised spatial DiD approach is given by

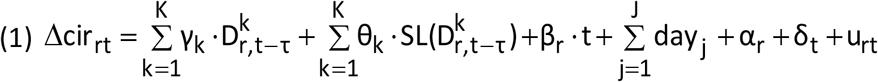

with

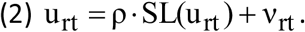

In the panel data model (1)-(2), the focus is directed on the response coefficients γ_k_ and θ_k_ of policy variables 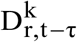 and their spatial lag 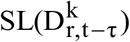, respectively. In view of an incubation period and a reporting delay (Linton et al., 2020; Lauer et al., 2020; Mitze et al. 2020), the treatment variables enter the model with a temporal lag of τ days. As the time index t refers to days, no observable time-varying regional traits are available as explanatory variables. They are captured by a spatially autocorrelated error process, where 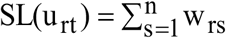 is the spatial lag in the disturbances u_rt_. ρ is the spatially autoregressive parameter of the error process. Time-constant variables like regional demopgraphic characteristics (e.g. population density, share of elderly population, average skill level of regional labor force) that are expected to affect the risk of infection, are captured by regional fixed effects. Finally, confounding factors common to all regions, such as national announcements by Federal politicians or changes in international travel restrictions by the German Foreign Office, are captured by time fixed effects.

We adopt a spatial error process from the start to prevent a potential omitted variable bias in absence of additional time-varying regressors at the level of daily data. Therefore, not the disturbances u_rt_ but the innovations ν_rt_ are assumed to be independently identically normally distributed. Equations (1)-(2) represent a spatial Durbin error model (SDEM) (Elhorst 2014, 10-12, 37-39; Bouayad-Agad, Le Gallo and Védrine 2018). The SDEM nests two simpler spatial models. Without policy spillovers, θ_k_=0, the spatial error model (SEM) is obtained. When other time-variant variables do not matter, ρ=0, the disturbances u_rt_ become white noise (u_rt_= ν_rt_). In this case, the SDEM coincides with SLX panel data model (Vega and Elhorst 2015).

To identify treatment effects for spatial data, Delgado and Florax (2015) develop a two-period difference-in-differences (DiD) model with local interregional interaction. Chagas, Azzoni and Almeida (2016) generalize the spatial DiD approach with respect to the type of interactions and number of time periods. They show that the average treatment effect on the treated (ATET) in an SLX-type model depends on the response coefficients of the treatment variable and its spatial lag as well as the average proportion of treated neighbours. As the error structure does not add an additional element, the ATET in the superordinate SDEM model is identically structured:

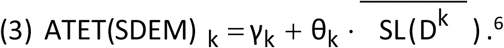

The treatment effect in (3) refers to the k*th* policy action. Non-treated regions benefit as well from interventions in nearby areas by spillover effects that amount to 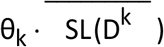. When the regions are consecutively included into the program, the ATET(SDEM) _k_ ultimately reaches the value γ_k_+ θ_k_ as the average share of adjacent treated regions, · 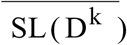, approaches 1. Because of disregarding local treatment effects, the standard DiD estimator is biased and inconsistent (Delgado and Florax 2015). In the spatial error model (SEM), the average treatment effect reduces to ATET(SEM)_k_ = γ_k_.

In view of the worldwide spreading of the virus, the spatial model (1)-(2) is still incomplete. People infect other people living within their residential areas and across administrative borders. While the propagation of infection is spatially global in nature, a distance decay from the origins is expected. Such global infection spillovers of Covid-19 can be captured by an endogenous spatial lag, 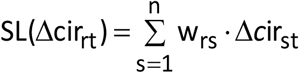, as an additional explanatory variable in the spatial panel data model (1):^7^

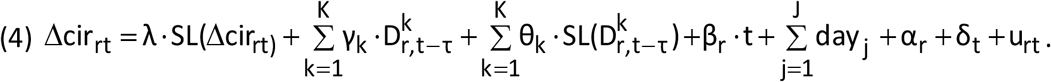

Equation (4) is known as the spatial Durbin model (SDM) (LeSage and Pace 2009, 32-36; Ehlhorst 2014, 37-38). If the disturbances u_rt_ are generated by the spatial autocorrelation process (2), equation (4) would represent the general nested spatial (GNS) model (Elhorst 2014, 37-38). However, because of its overparameterization and weak identifiability (Elhorst 2014, 32-39; Floch Le Saout 2018, 154-155), the GNS model is considered of minor empirical relevance.^8^ The spatial lag SL(Δcir_rt_) reflects the average log difference of cumulative infections per 100,000 inhabitants in the bordering areas of region r. Its regression coefficient λ is the spatial autoregressive parameter. While the within estimator is consistent with fixed T for n→∞ in the SLX model (1) (cf. Pesaran 2015, 640-644), this property gets lost in the spatial Durbin model in virtue of the presence of the endogenous spatial lag. To eliminate the simultaneity bias of the fixed effects (FE) estimator, the spatial panel data model (4) is estimated by maximum likelihood techniques (Lee and Yu, 2010).

Having accounted for the SUTVA and parallel trends assumption in the modelling approach for identifying causal effects of the public health measures, light needs to be shed on the issue of exogeneity. The use of the policy variables with temporal lags itself does not resolve the problem. Reverse causation will be present when local authorities introduce the measures, for instance, in response to increasing regional infection rates (Wooldridge 2010, p. 321). Here, however, actions are taken at a higher regional level in the framework of the Federal-State-Agreement (BLV). The regulations are then set in force by the states partially staggered in time.^9^ The implementation of public health measures at a subordinate level backs the validity of the exogeneity assumption as it can be argued that they do not account for the specific local differences (Carpenter and Dobkin 2011; Angrist and Pischke 2015).

In the presence of endogenous spillovers, the treatment effects are influenced by feedback effects that are not reflected in the response coefficients. Using matrix representation it can be shown that their influence is captured by the inverse matrix (**I** − λ · **W**)^−1^ with **I** as an nxn identity matrix. In the spatial Durbin model (4), impacts involving spillover effects are given by the product of the matrices (**I** − λ · **W**)^−1^and (γ ·**I** + θ· **W**) at each point in time (LeSage and Pace 2009, 34-42; Elhorst 2014, 20-23):

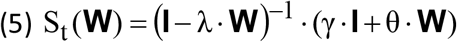

The infinite series expansion of the matrix (**I** − λ · **W**)^−1^,

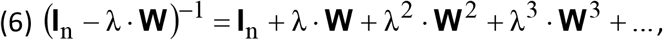

allows a decomposition of the impacts on first-order neighbours (**W**), second-order neighbours (**W**^2^), …, q*th*-order neighbours (**W**^q^) after which the terms on the right-hand side with |λ | < 1 become virtually zero.

By employing (5) at each point in time, different average impact measures can easily be computed. For a row-standardised weights matrix, the average total impact amounts to (γ + θ)/(1-λ). In accordance with (3), we can account for the share of treated neighbouring regions, 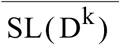, to obtain the average treatment effect on the treated (ATET) for policy action k in the spatial Durbin model (4):

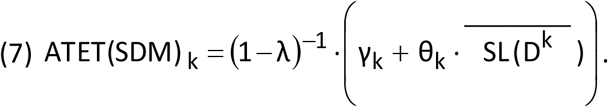

As regions are included into the treatment successively, ATET(SDM)_k_ eventually amounts to (γ_k_ + θ_k_)/(1-λ).

In estimating the spatial panel data models (1)/(2), (4) and (4)/(2), we have to make allowance for the fact that multicollinearity is provoked by a high correlation between the policy variable 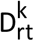 and its spatial lag 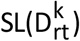. While the combined effect of the own and adjacent regions, γ_k_ + θ_k_, can be precisely estimated, its partitioning becomes more and more arbitrary with increasing strength of correlation. This is particularly the case here where the panel data analysis covers a large number of points in time at which the regions are consecutively included into the intervention. Hence, first we estimate the combined policy effects γ_k_ + θ_k_ in the SDEM and SDM model. In the latter model, we thereafter identify the total treatment effects (7) by impact analysis. Finally, a spatial partitioning of the total impacts with respect to their feedback effects is accomplished. Significance testing is performed by Monte Carlo simulation (Piras 2013).

## 5 Empirical Evidence

In adopting the spatial difference-in-difference approach, causal effects of public health measures to contain the spread of Covid-19 are worked out. According to our quasi-experimental design, we aim to identify incremental impacts of the different categories of containment measures on the spread of Covid-19 until late April. In order to control for the pre-treatment development, the sample period starts on February 24, 2020. Prior to that time, only occasional infection cases were reported to local public health authorities. In order to avoid overlapping with easing measures, the period of investigation ends on April 24, 2020.

The Space–Time Moran’s I (STMI) of 0.3223 (p<0.001) reveals that infection rates are highly significantly spatially autocorrelated across German NUTS-3 regions. The presence of spatial spillovers in the spreading of Covid-19 is a breach of the Stable Unit Treatment Value Assumption (SUTVA) on which the usual DiD approach is based. Thus, we make use of the spatial DiD approach that explicitly allows for interactions between regional populations in estimating impacts of the containment measures. Furthermore, the parallel trend assumption is addressed by the including region-specific trends. With only some exceptions, the estimated trend coefficients are statistically significant at least at the 5% level.

Because of peculiarities in reporting behavior over the week, the day of the week effect is included in spatial regressions. Strong differences between days illustrate the necessity to adjust for daily seasonality (Figure 8, Appendix C). Highly significant deviations from the reference day (Sunday) are observed for all days. Particularly because of limited testing on weekends, figures begin to rise at the start of the week and fall after a peak in the middle of the week on Thursday. The maximum difference amounts to almost 5 percentage points.

The econometric estimation of spatial panel data models is the first step in identifying causal effects of the spatial DiD approach. To account for incubation time and reporting delay, the policy dummies are introduced into the model with a one-week lag.^10^ In this step, the SEM and SDEM models are estimated as spatial baseline models to check robustness and to test the common factor hypothesis. The maximum likelihood (ML) estimates of the response coefficients and the spatial autoregressive parameter of the SDM model are used in a second step to establish the feedback and total effects by impact analysis. Because the implementation of a measure is closely staggered in time across the regions, the treatment variable tends to be highly correlated with its spatial lag. While the combined effect of each policy action is estimated correctly, the attribution of influences from the own and neighbouring regions becomes arbitrary. We therefore focus on the combined effect that is uniformly reported for all estimated panel models.

In accordance to the STMI test, spatial spillovers show up in spatial autoregressive process of infection growth in all models. Although the spatial autoregressive has a different interpretation in the SEM and SDEM model, its estimate is in all panel data models close to 0.08 (Table 2). The relatively low value presumably reflects restrictions in spatial interaction like job commuting and travelling across regions during the period of investigation. However, the estimates are highly significant. The necessitiy to incorporate spatial effects is strongly corroborated by Pesaran’s local CD test and Breusch and Pagan’s local scaled LM test.

**Table 1:** Summary statistics

| Variable | Mean | Median | Standard deviation | Minimum | Maximum |
| --- | --- | --- | --- | --- | --- |
| *Raw data on number of cases:* |  |  |  |  |  |
| New Covid-19 cases | 6.33 | 2.00 | 13.99 | 0 | 311 |
| Cumulative Covid-19 cases | 149.50 | 44.00 | 332.20 | 0 | 5600 |
| *Raw data on case incidence:* |  |  |  |  |  |
| Covid-19 incidence rates | 3.09 | 1.15 | 5.44 | 0 | 142.06 |
| Cumulative Covid-19 incidence rates | 71.26 | 31.35 | 106.26 | 0 | 1517.16 |
| *Dependent variable:* |  |  |  |  |  |
| Cumulative incidence growth rate | 0.089 | 0.018 | 0.188 | 0 | 2.833 |
| *Containment measures:* |  |  |  |  |  |
| Establishment closure | 0.63 | 1 | 0.48 | 0 | 1 |
| School closure | 0.65 | 1 | 0.48 | 0 | 1 |
| Shopping mall closure | 0.56 | 1 | 0.50 | 0 | 1 |
| Restaurant closure | 0.57 | 1 | 0.50 | 0 | 1 |
| Contact restrictions | 0.54 | 1 | 0.50 | 0 | 1 |
| Mask duty | 0.01 | 0 | 0.08 | 0 | 1 |
| *Delays between infection / first day of symptoms and reporting* |  |  |  |  |  |
| Infection and reporting | 11.7 | 10.5 | 6.05 | 3.5 | 34 |
| First symptoms and reporting | 5.5 | 4.0 | 5.72 | 0 | 26 |

*Note:* Raw data on number of cases comes from RKI (2020a). We conduct our analysis on the NUTS-3 level with 401 regions. Our analysis starts on February 24 and ends on April 24, 2020, which results in 61 days and 24,451 observations in total. Incidence rates are defined by Covid-19 cases per 100,000 inhabitants. We report the descriptive statistics for the containment measures without time lag. Summary statistics on the delay between first day of symptoms and the day of reporting (reporting lag) are computed from RKI (2020a) database. Summary statistics on delay between infection and the day of reporting (overall lag) come from Mitze et al. (2020). For both delays the extreme values are given by the 1st and 99th percentiles.

**Table 2:**
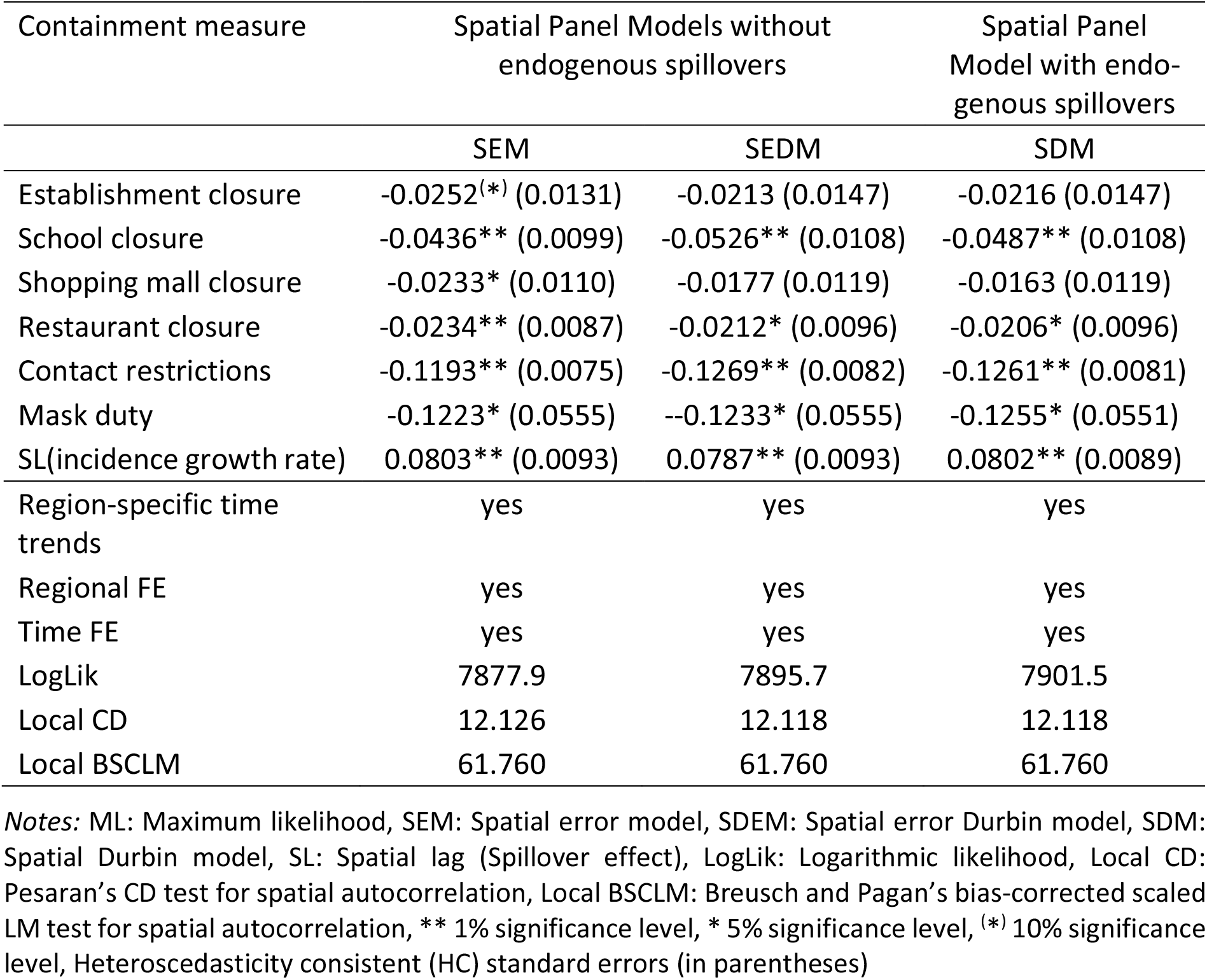
ML estimated spatial panel data models

While all estimated response coefficients of the six compound public health measures show the expected negative sign, not all are statistically significant (Table 2). Substantial effects of the closure of establishment and the shutdown of non-essential retail stores in shopping malls and elsewhere suggested by the baseline SEM model are not confirmed by the spatial models with intervention spillovers. This indicates an omitted variable bias in the SEM model. As the common factor hypothesis is clearly rejected (LR=47.2, p<0.001), its interpretation as a simplified SDM model also fails. Because of LogLik(SDM) > LogLik(SDEM) with the same number of explanatory variables, information criteria AIC and BIC select the SDM model as the optimal setting.

We therefore focus on the estimated regression coefficients of the SDM model that reflect the raw effect sizes of the policy interventions in the own and surrounding areas. Feedback effects are neglected at this step. With estimated values of about -0.125, the absolutely largest sizes are found for the implementation of contact restrictions^11^ and mandatory face mask wearing in public transport and sales shops. The lower statistical significance of the estimated face mask effect results from the small number of treated areas in the containment phase. A smaller but still strong response coefficient of nearly -0.05 is estimated for closure of schools, further educational institutions and daycare facilities. As in the case of contact restrictions, the estimated regression coefficient of school closure is highly significant. An incremental containment effect may possibly also be attributed to the closure of restaurants, bistros, pubs, bars and cafés. However, the value of -0.02 indicates at best a limited impact of this policy action.

In the second step, we augment the raw effects of the public health measures by feedback effects. People may infect inhabitants in surrounding areas who − in turn − infect residents in the region of origin (and *vice versa*). Total treatment effects and their composition can be obtained by impact analysis (LeSage and Pace 2010, 366-372). In Table 5 impact estimates are reported for the SDM model with policy actions according to state regulations. The spatial DiD approach clearly reveals the relevance and strength of public health measures to curb the pandemic. As suggested by ML estimation of spatial panel data models, the highest impacts arise from contact restrictions and face mask wearing. Effects from school closure follow with a clear gap in terms of effect size. The closure of restaurants may additionally exert a marginal impact on infection growth. Because of the inclusion of feedback effects, the total effects are nearly 10 percent larger than the response coefficients.

Having witnessed daily growth of incidence rates of more than 50 percent in the pre-treatment period, the dampening effect of contact restrictions is estimated by almost 14 percentage points. The total treatment effect of school closure amounts to 5½ percentage points. However, as a ban on major events took place around the same time as the school closures, the estimated effect from this type of containment measure might potentially be upwardly biased. Both impacts turn out to be highly significant. With 2¼ percentage points, the effect of restaurant closure appears to be relatively moderate. At the end of the containment phase, a disease mitigating effect of mandatory face mask wearing is found. Contrary to the other measures, the strong impact of 13½ percentage points (on the cumulative incidence growth rate) only relates to the few treated regions during our sample period. This explains its relatively weak statistical significance. No significant treatment effects are found for closure of establishments and the shutdown of shopping malls and other non-essential retail stores. While their impacts exhibit the expected sign, no substantial contribution of both policy actions to flatten the infection curve is apparent.

Given the estimated response coefficients of the SDM model (Table 2), the impacts are not uniform and vary across space (LeSage and Pace 2009, 33-39). In Table 3 concise summary measures of impacts are reported. As average values they are representative for the whole area but not necessarily for each region individually. The density plots of in Figure 2 show the distributions of the total impacts from the SDM model. They provide particularly an impression on their variability across space. While the effects of contact restrictions and school closure are precisely estimated, the impact of face masks varies more broadly. This lower degree of estimation precision results from a comparatively large sample error as a consequence of a small number of treatment regions during the containment period. More generally, the density plots provide information on the extent of spatial non-stationarity that is involved with the implementation of the public health measures.

**Table 3:** Impact decomposition of containment measures

|  | Combined localised impacts | Feedback impacts | Total impacts |
| --- | --- | --- | --- |
|  | SDM panel model |  |  |
| Establishment closure | -0.0216 (0.0148) | -0.0019 (0.0013) | -0.0235 (0.0161) |
| School closure | -0.0487** (0.0102) | -0.0042** (0.0010) | -0.0529** (0.0111) |
| Shopping mall closure | -0.0163 (0.0116) | -0.0010 (0.0012) | -0.0177 (0.0126) |
| Restaurant closure | -0.0206* (0.0098) | -0.0018* (0.0009) | -0.0224* (0.0106) |
| Contact restrictions | -0.1261** (0.0082) | -0.0110** (0.0015) | -0.1371** (0.0090) |
| Mask duty | -0.1255* (0.0551) | -0.0109* (0.0051) | -0.1364* (0.0599) |

<sup>11</sup> If we assume that contact restrictions were also effective in Rhineland-Palatinate and Thuringa the day following their announcement, estimated response coefficient of this policy action slightly drops to 0.12 in absolute terms. Thus, a stronger impact of this measure on growth rates of infections is associated with its actual implementation.
Notes: Simulated standard errors (in parentheses): Monte Carlo simulation (Piras 2013), \*\* 1% significance level, \* 5% significance level, (\*) 10% significance level

**Table 5:** Implementation dates of containment measures at federal state level

| Federal state | # NUTS-3 districts | Establishment closure | School closure | Shopping mall closure | Restaurant closure | Contact restrictions | Mask duty |
| --- | --- | --- | --- | --- | --- | --- | --- |
| Baden-Wuerttemberg | 44 | 17.03. | 17.03. | 18.03. | 21.03. | 23.03. | 27.04. |
| Bavaria | 96 | 17.03. | 16.03. | 18.03. | 21.03. | 21.03. | 27.04. |
| Berlin | 1 | 15.03. | 17.03. | 23.03. | 22.03. | 23.03. | 27.04. |
| Brandenburg | 18 | 18.03. | 18.03. | 18.03. | 23.03. | 23.03. | 27.04. |
| Bremen | 2 | 18.03. | 16.03. | 18.03. | 21.03. | 23.03. | 27.04. |
| Hamburg | 1 | 16.03. | 16.03. | 17.03. | 21.03. | 23.03. | 27.04. |
| Hesse | 26 | 18.03. | 16.03. | 18.03. | 21.03. | 23.03. | 27.04. |
| Lower Saxony | 45 | 17.03. | 16.03. | 17.03. | 20.03. | 23.03. | 27.04. |
| Mecklenburg-West P. | 8 | 18.03. | 16.03. | 18.03. | 21.03. | 23.03. | 27.04. |
| North Rhine-Westph. | 53 | 16.03. | 16.03. | 18.03. | 23.03. | 23.03. | 27.04. |
| Rhineland-Palatinate | 36 | 18.03. | 16.03. | 18.03. | 21.03. | 23.03. | 27.04. |
| Saarland | 6 | 18.03. | 18.03. | 18.03. | 21.03. | 21.03. | 27.04. |
| Saxony | 13 | 19.03. | 18.03. | 19.03. | 21.03. | 23.03. | 20.04. |
| Saxony-Anhalt | 14 | 18.03. | 16.03. | 18.03. | 25.03. | 23.03. | 22.04. |
| Schleswig-Holstein | 15 | 15.03. | 16.03. | 18.03. | 18.03. | 23.03. | 29.04. |
| Thuringia | 23 | 18.03. | 17.03. | 20.03. | 20.03. | 23.03. | 24.04. |

*Note:* There are 16 federal states (NUTS-1 regions) in Germany. The federal states are sub-divided into 401 NUTS-3 districts. In most cases, the containment measures were put in place at the federal state level. If this is the case, the measure is binding for all NUTS-3 districts within the corresponding federal state. We did our best to check for different implementation dates in specific NUTS-3 districts. If we found differences, we account for them and state them in the main text.

**Figure 2:**
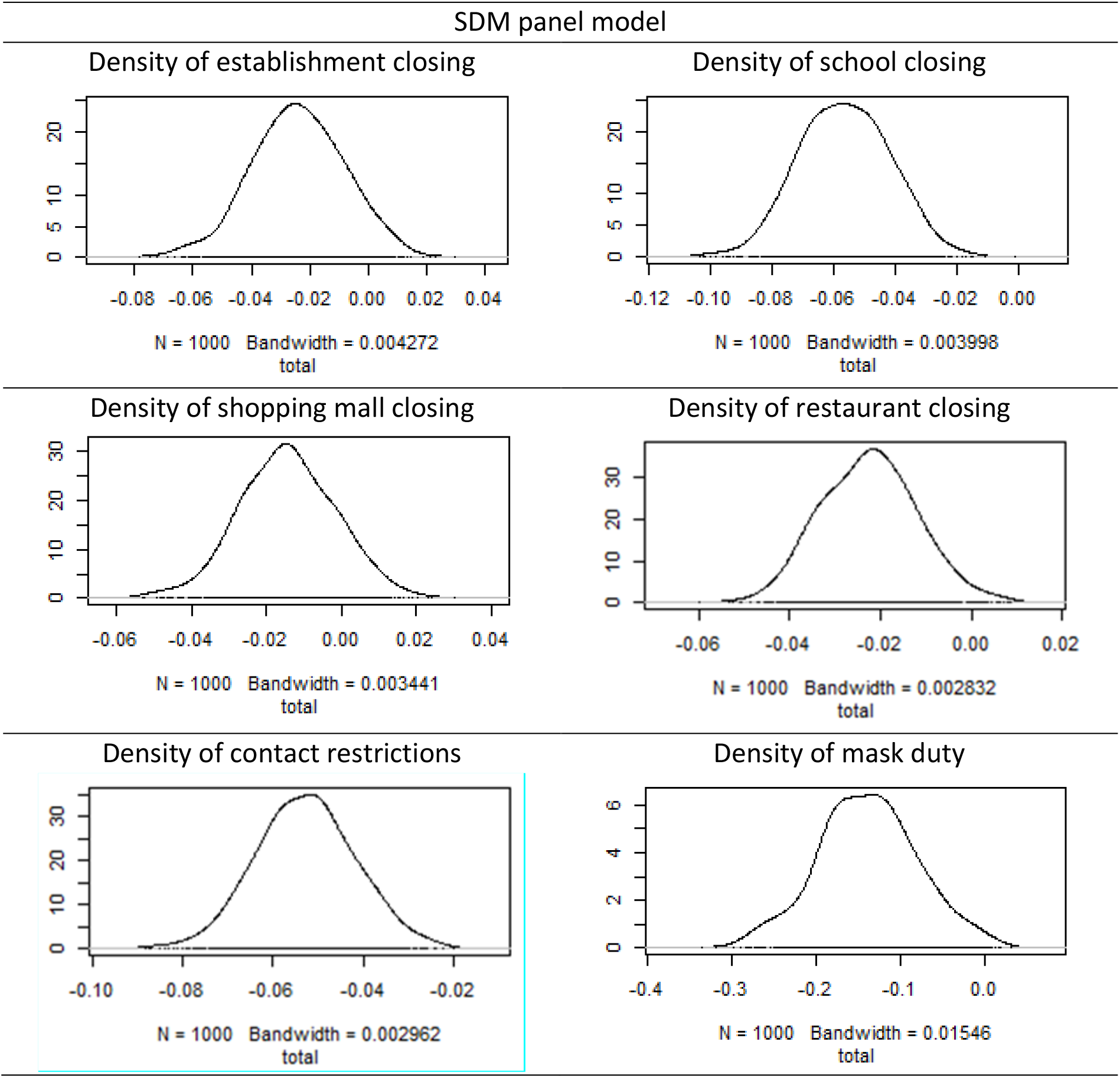
Density plots for total impacts of public health measures

Above we have estimated the treatment effects of the most important public health measures. Contact restrictions, face mask wearing and school closure are shown to yield substantial containment effects in treated regions. Additionally, a weak impact may be attributed to restaurant and further leisure facility closure. While the treatment effects of school and restaurant closure are established before other measures were enacted, the impact of the mask duty arises as an add-on effect. We now set our sights from infections in the treated regions to the pandemic curve in Germany as a whole. To show the overall containment effect of individual measures, we have constructed a counterfactual curve by aggregating regional containment effects at different points in time.

In Figure 3 the temporal impacts temporal impacts of the policy actions on the German pandemic curve are illustrated. For this purpose, the region-specific treatment effects are translated into reduction effects of the national pandemic curve. The comparison between the actual and counterfactual infection curve exposes the incremental contribution of a policy action to dampening the propagation of the virus. Each illustration starts with a pre-treatment period of one week. After the vertical blue line, the actual and counterfactual trajectories of cumulative disease cases are shown on the logarithmic scale for a two week treatment period. While the black lines display the real evolution of the pandemic, the blue lines reflect its counterfactual course without implementing the particular health measure. The red lines exhibit the 95% confidence bands.

**Figure 3:**
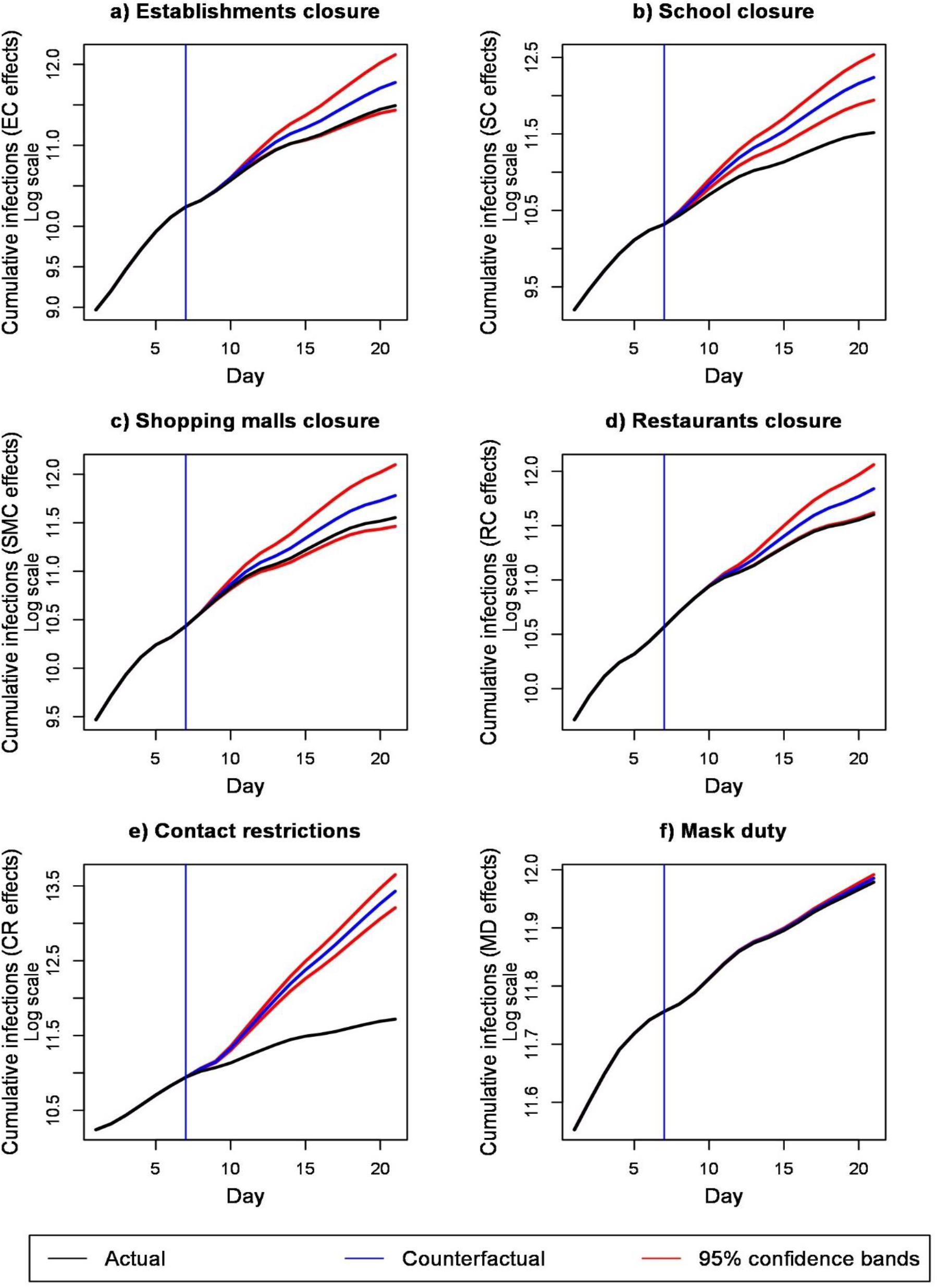
Temporal impacts of health measures on the German pandemic curve. *Notes:* Vertical blue line: Separation between pre-treatment period and treatment period; Length of pre-treatment period: one week; length of treatment period: two weeks

Figure 3 exhibits the substantial contributions of contact restrictions and closure of schools to flatten the pandemic curve. Although the confidence intervals widen with increasing total infection rates, the counterfactual curves and their significance bands of both measures lie well above the actual curve. While the containment effect of contact restrictions is sizable, the impact of school closure is still noticeable. However, the the observed speed of the flattening of the pandemic curve may point at the presence of announcement effects that led to early cancellations of large events. The latter would confirm that the estimated effects from closing schools and daycare facilities constitute an upper bound for the true effect from this type of public health intervention. The overall effect of the closure of restaurants is markedly smaller. Its lower 95% confidence band is only just above the observed infection trajectory. The visibility of the impacts at different points in time after the one-week delay is mainly due to the fact that the interventions are not uniformly implemented staggered in time. Policy-specific aspects like announcement effects may additionally play a role.

Although the treatment effect for face mask wearing is strong, this policy action appears to only marginally affect the national infection curve. The obvious reason for this is that we analyze impacts in the containment phase where the face mask duty is only implemented in very few regions. Its cumulative effect on the German pandemic curve can thus hardly be compared to the effect size of the other measures, which have been eventually implemented in all NUTS-3 districts during our sample period. Accordingly, while we establish a strong treatment effect in these regions, the summed impact on cumulative infections in the whole area is small. This would surely change if more regions were included in the treatment group – conditional on the fact that the estimated coefficient would remain statistically significant and negative. However, a nationwide face mask mandate became only effective after our sample period had ended. For establishment closing and the shutdown of shopping malls and other stores, the actual infection curve runs inside the confidence areas of the counterfactual curves. This behaviour reflects the estimated non-significant treatment effects of both policy actions.

## 6 Robustness

In sensitivity analyses we check the robustness of containment effects of public health measures with respect to various changes in modeling design. We already showed the robustness of the treatment effects of the SDM model against baseline fixed-effects models with spatially autocorrelated disturbances. Here, we first analyze the robustness of the findings as related to variations of the spatial DiD approach with endogenous spillovers. Two alternative spatial panel data models appear to be of special interest. One is the spatial autoregressive (SAR) model that makes allowance for endogenous spatial spillovers but does not explicitly account for policy interventions in neighbouring regions.^12^ The other is the general nesting spatial (GNS) model that allows for spatial spillovers in the endogenous variables, the policy variables and the disturbances.

Figure 4 shows limited variations between the effect estimates of the three spatial models. The estimated response coefficients of contact restrictions, wearing of face mask, school closure and restaurant closure are statistically significant in all models. While weak significant effects of establishment and shopping mall closure is suggested by the SAR model (blue bars), the nonsignificance of both containment measures is corroborated by the GNS model (red bars). However, the problems of overparameterization and weakly identifiability of the GNS model becomes evident (Elhorst 2014, 32-33). Estimates of the spatial autoregressive coefficients for endogenous spillovers (0.1721) and error dependence (-0.1038) are blown up with opposing signs (Elhorst 2017). Along with the fact that only the difference of the spillover parameters conforms with the estimates of more parsimonious spatial representations, this makes the empirical relevance of the GNS model highly doubtful.

**Figure 4:**
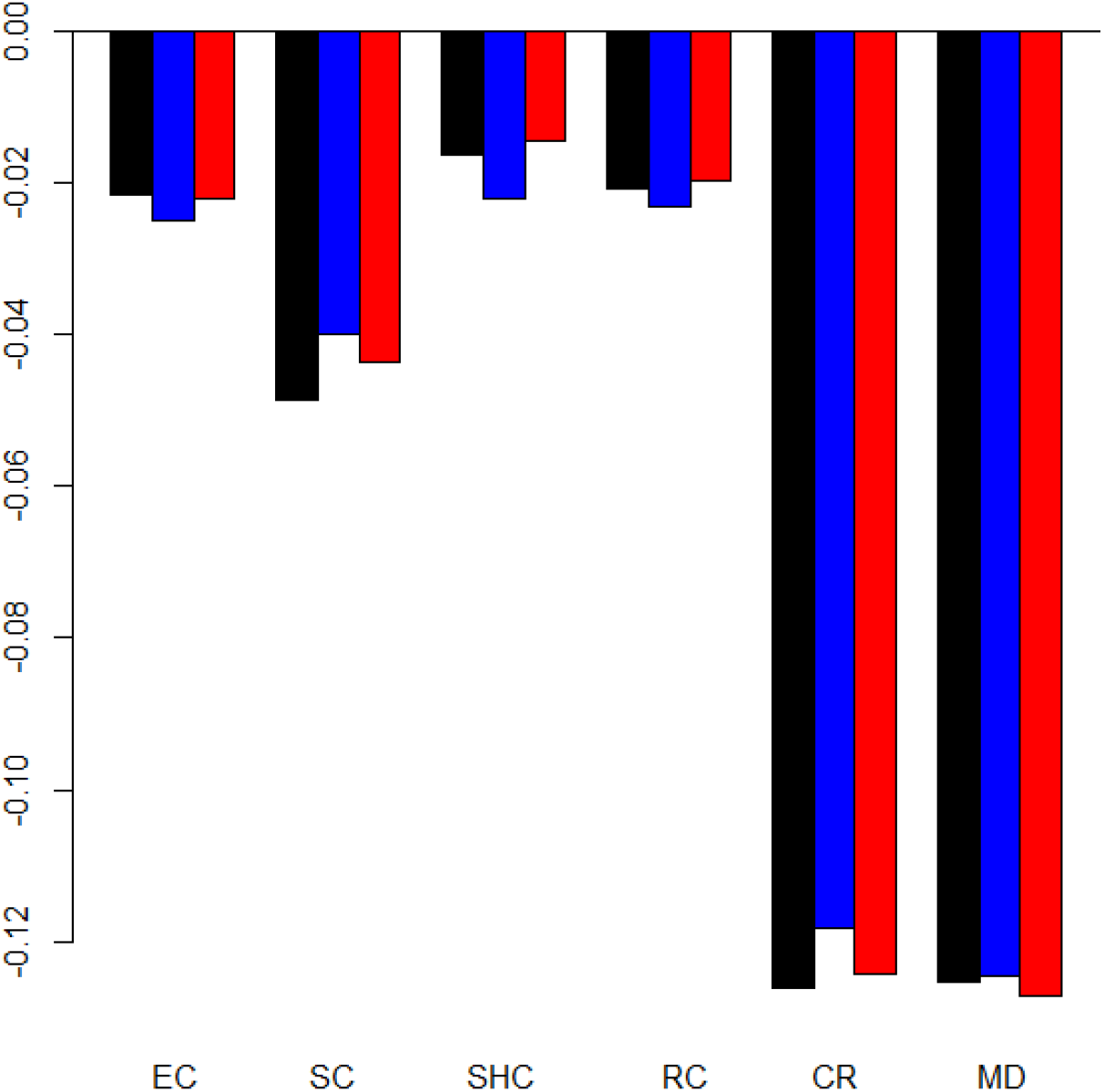
Comparison of response coefficients of SDM, SAR and GNS models. *Notes:* Horizontal axis: Public health measures, vertical axis: Response coefficients of spatial panel data models, Black bars: SDM model, blue bars: SAR model, red bars: GNS model, EC: Establishment closure, SC: School and daycare facilities closure, SHC: Shopping mall closure, RC: Restaurant closure, CR: Contact restrictions, MD: Mask duty

Next, we examine the robustness of treatment effects with respect to the choice of the temporal lag for the included containment measures. In view of the incubation time and reporting delay, we expect that after about one week one fourth of new infections are reflected in registered cases (Mitze et al. 2020). According to the joint distribution of both lags, official figures will capture different shares of infected people with varying delays. If containment effects are substantial, they should not only be observed at a unique lag. Thus, we check the effects for some days back and ahead of the reference lag of one week. In this context, we also review the stability of the estimated spatial spillover coefficient.

The outcomes of the robustness checks on lag variation are depicted in Figure 5. The delay between the outbreak and the report of infections is varied between 5 and 10 days.^13^ The left panel (Figure 5a) shows the high stability of estimated response coefficients. This holds especially for the effects of contact restrictions and mandatory face mask wearing. Statistical significance of school closure and restaurant closure is most notably confirmed with increased delays. For school closure, significance comes along with an intensified treatment effect. The deviations for the lag of 5 days may be due to the short interval for identifying impacts arising from this specific measure. In the right panel (Figure 5b) a slight tendency of decreasing spillover coefficients is apparent with increasing lag. However, all estimates are around 0.08 and their differences remain within reasonable limits.

**Figure 5:**
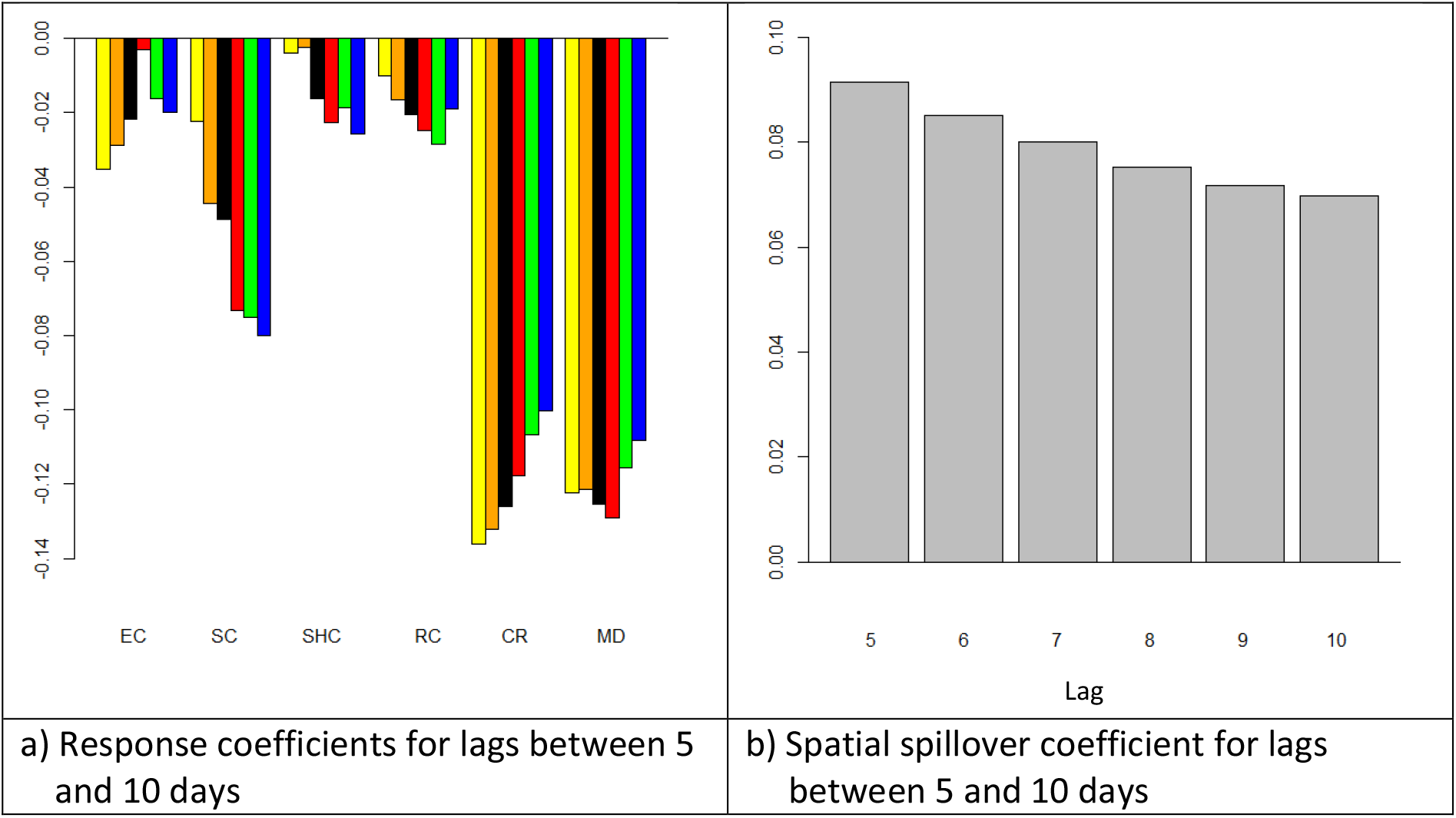
Comparison of response and spillover coefficients for varying lags. Notes: Horizontal axis: Public health measures, vertical axis: Response coefficients of spatial panel data models, Yellow bars: Lag 5, Orange bars: Lag 6, Black bars: Lag 7, Red bars: Lag 8, Green bars. Lag 9, Blue bars: Lag 10, EC: Establishment closure, SC: School and daycare facilities closure, SHC: Shopping mall closure, RC: Restaurant closure, CR: Contact restrictions, MD: Mask duty

The dashboard data of the Robert Koch Institute (RKI) on Covid-19 cases reflect some anomalies of reporting over the week. Generally, infection cases are less reported on weekends and holidays than on workdays. In our modeling approach we capture cyclicity over the week by a day-of-the-week factor. Through this, we also account for erratic reporting of with low and large numbers of newly confirmed cases occurring in immediate succession. One can alternatively control for intra-week seasonalities by smoothing the data. To check the robustness of treatment effects to modeling the cyclical reporting pattern, we re-estimate the SDM model with three and five-days moving averages of incidence rates.

Figure 6 compares the estimation results for the original and smoothed infection data. It is evident that the response coefficients of public health measure are not substantially affected by the filtering procedure. With increasing order of the moving average, the nonsignificant regression coefficient of shopping mall closure approaches zero. Also, the estimated effects of contact restrictions and face mask wearing decrease, but without losing their relative strength. However, filtering newly registered cases is a data transformation that changes their dispersion. Thus, averaging the original data has expected attenuation effects on the standard errors. For the 3-days and 5-days moving averages the standard errors of the estimated regression coefficients are reduced by factors of about 1.6 and 2, respectively. This impact, that favors the detection of treatment effects, is not considered by Brauner et al. (2020), who establish the effectiveness of non-pharmaceutical interventions in 41 countries on the basis of smoothed death and cases. To avoid a dependence of testing results on the order of moving averages, we rely final significance testing on the original data.

**Figure 6:**
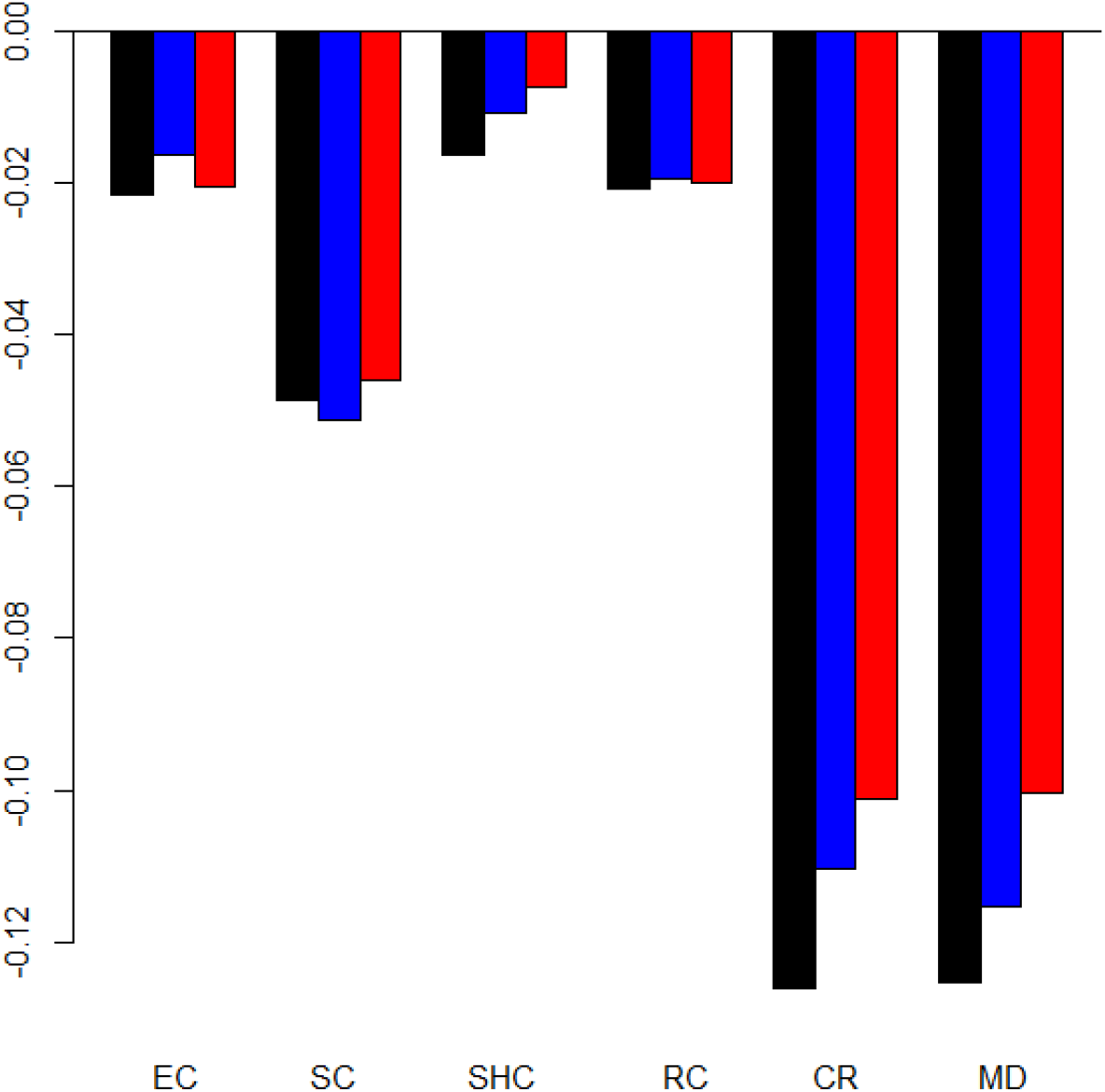
Smoothing effects on response coefficients. *Notes:* Horizontal axis: Public health measures, vertical axis: Response coefficients of spatial panel data models, Black bars: SDM model with original data, blue bars: SDM model with three days moving averages, red bars: GDM model with five days moving averages, EC: Establishment closure, SC: School and daycare facilities closure, SHC: Shopping mall closure, RC: Restaurant closure, CR: Contact restrictions, MD: Mask duty

Finally, we assess the sensitivity of the response coefficients with respect to the choice of the spatial weights matrix. Figure 7 displays a comparison of the policy effects obtained from ML estimation of the SDM model with the contiguity- and distance-based weights matrices. In the latter case, we make use of inverse distances with cut-off thresholds of 75 km, 100 km and 150 km. For comparative purposes, all spatial weights matrices are row-standardized.

**Figure 7:**
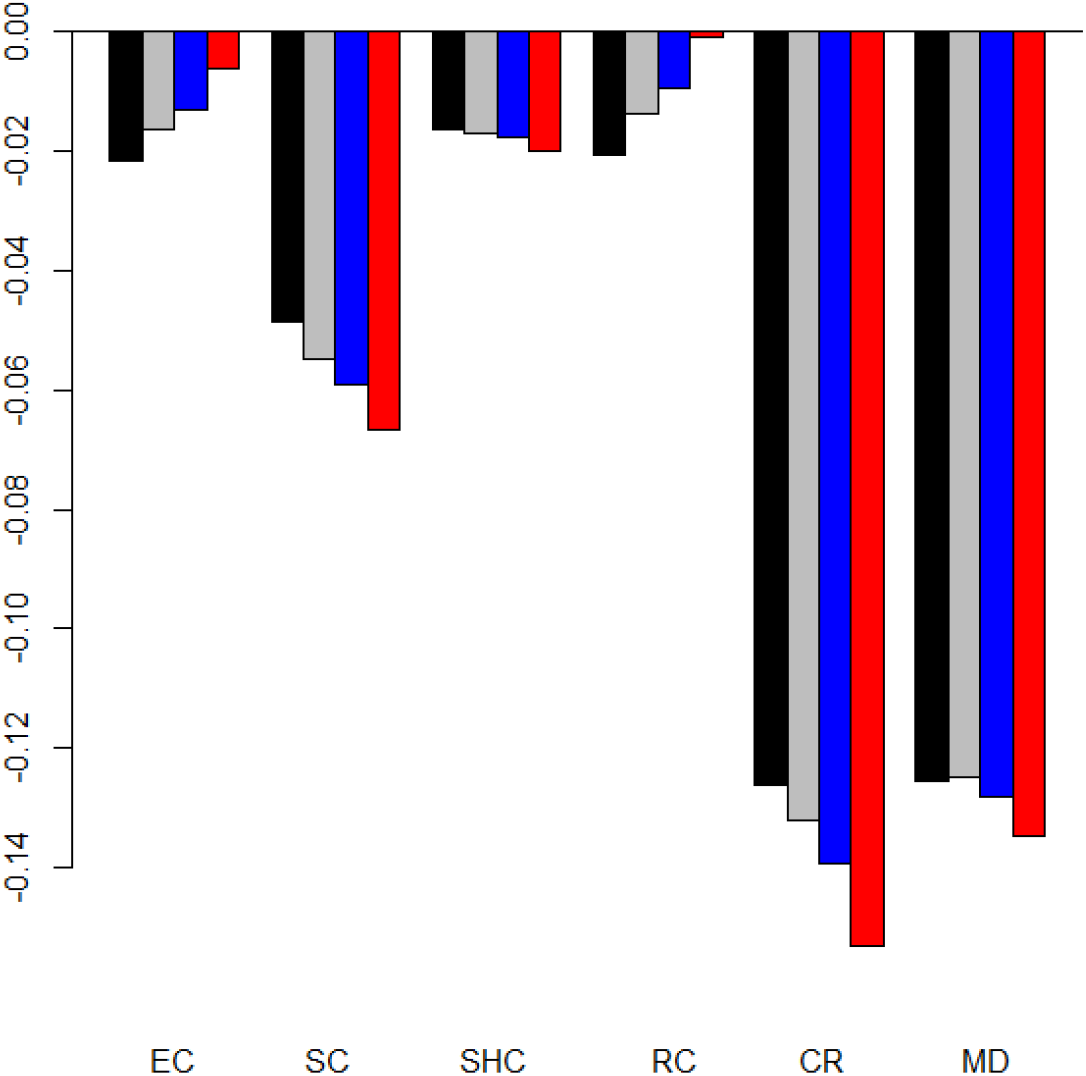
Comparison of response coefficients for different spatial weights matrices. *Notes:* Horizontal axis: Public health measures, vertical axis: Response coefficients of spatial panel data models, Black bars: Contiguity weights matrix, Grey bars: Inverse distance weights matrix (cut-off threshold: 75 km), Blue bars: Inverse distance weights matrix (cut-off threshold: 100 km), Red bars: Inverse distance weights matrix (cut-off threshold: 150 km), EC: Establishment closure, SC: School and daycare facilities closure, SHC: Shopping mall closure, RC: Restaurant closure, CR: Contact restrictions, MD: Mask duty

**Figure 8:**
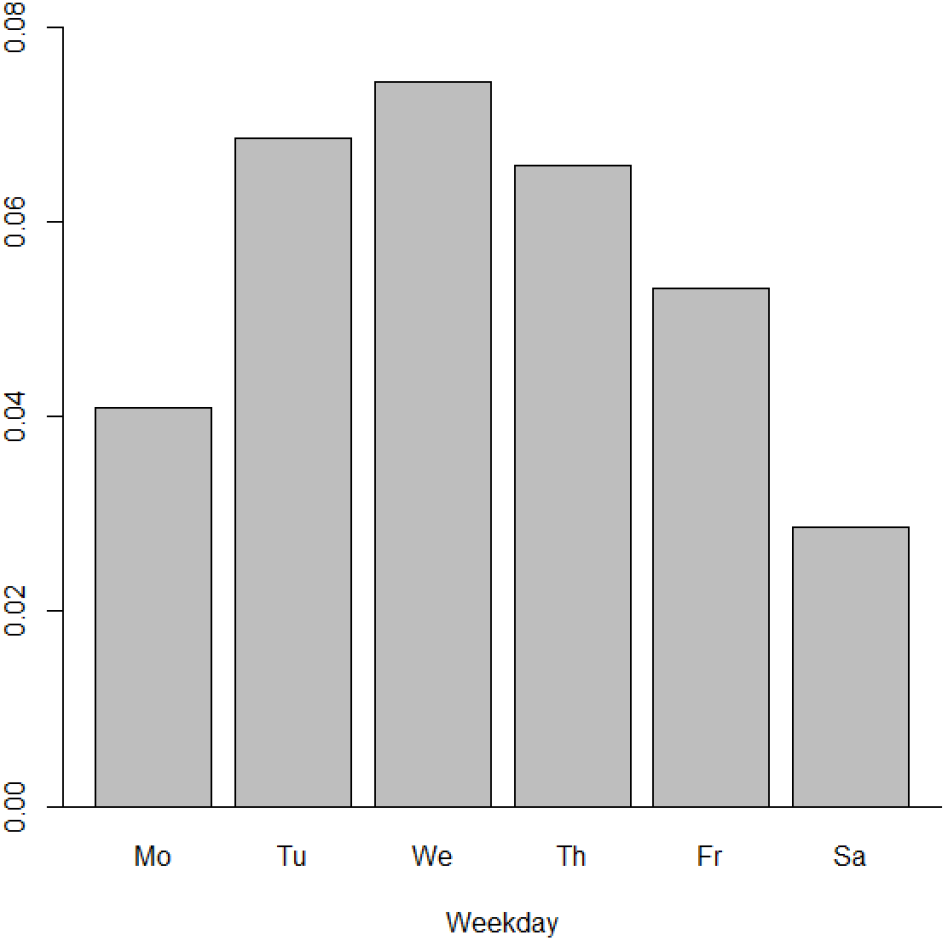
Day of the week effect

Figure 7 exhibits a high degree of robustness of estimated policy impacts across all spatial weights matrices. Regardless of the selected type of weights matrix, contacts restrictions prove to be the most effective measure to curb the pandemic development during spring 2020. The strong treatment effect of mandatory face mask wearing is clearly confirmed with distance-based weights matrices. Sizable infection effects of closure of schools and daycare facilities hold as well independently of the employed spatial weights concept. On the contrary, the significant impact of restaurant closure on infection growth is not backed with distance-based weights matrices. Sensitivity analysis corroborates the lack of a substantial influence of establishment and non-essential retail store closure on the German pandemic curve.

## 7 Conclusions

We have tackled the challenge of analyzing the impacts of public health measures to curb the spread of Covid-19 in Germany. In view of the constrains to identify causal effects in the case of incremental implementation of these non-pharmaceutical interventions, we have assessed six types of compound measures. After the closure of educational institutions including schools and daycare facilities as well as a range of establishments as of mid-March, non-essential retail stores in shopping malls and other locations, restaurants, bars and other gastronomic facilities were shut down. Strict contact restrictions have been introduced in the second half of March. During the containment phase, the wearing of face masks (in public transportation and sales shops) has become mandatory in some NUTS 3 regions before the regulation has been implemented by all state governments. Parallel to this, other restrictions have been gradually lifted to allow for a re-opening of the economy.

Our identification strategy takes up temporal changes and regional differences of implemented regulations. The spatial differences-in-differences (DiD) model exploits the variations of the policy actions to identify their mitigating effects on the cumulative incidence gowth rate in the 401 German NUTS-3 regions. In the quasi-experimental approach, we involve region-specific trends to account for the parallel trend assumption. We are convinced that the study design is adequate to identify causal effects of the public health measures. The spatial econometric framework allows for inter-regional policy effects as well as endogenous spatial infections spillovers. We control for omitted time-invariant variables and nationwide infection trends. Finally, we are aware of the necessity to control for pronounced day-of-the-week anomalies in identifying the treatment effects.

Various tests on the existence of spatial spillovers clearly back the spatial econometric approach chosen here. In the presence of endogenous infection spillovers, we have to account for feedback effects to adequately identify treatment effects of policy interventions. On that score, the response coefficients of the spatial Durbin model have been re-assessed by impact analysis. We found the greatest containment effect of nearly 14 percentage points (on the cumulative incidence growth rate) for contact restrictions, followed by wearing of face masks with about 13½ percentage points. While the impact of contact restrictions is grounded on comprehensive regional implementations during the period of study, that of face mask wearing is only based on a few treated regions during the covered containment phase. But both measures are thus effective in flattening the German pandemic curve. However, the more variable (and in absolute terms smaller) impact of mask wearing is a consequence of our sample design to not confound the effects from containment and easing measures. On the basis of a DiD approach, Deb et al. (2020) confirm that contact restrictions brought about a significant reduction of the growth of Covid-19 cases in a 30-day period. The effectivesness of mandatory face mask wearing is corroborated by Mitze et al. (2020).

We also find evidence for a substantial influence of the closure of schools and daycare facilities in suppressing the spread of Covid-19. The statistically significant impact amounts to almost 5½ percentage points. This estimated effect might, though, reflect an upper bound of the true effect of school closure as the ban of major events (not explicitly covered in our analysis) took place around the same time. However, in an international comparison, Deb et al. (2020) found an even stronger impact of school closures on the pandemic curve. Persson, Parie and Feuerriegel (2021) find evidence of a containment effect of school closures for Switzerland.^14^

While the impacts of contact restrictions, face mask wearing and school closure prove to be robust against changes in model specification, the relatively smaller containment effect of about 2 percentage points for closure of restaurants and other gastronomic facilities does not emerge for all variations of the modeling approach. No significant effects to curb the growth of infections are found for the shutdown of shopping malls and other stores as well as the closure of establishments. This finding is shown to be robust in our sensitivity analysis. Spatial spillovers account for almost 10 percent of the cumulative incidence growth rate at the regional level. Because spatial dependence is found to be a relevant determinant of the local infection dynamics, a strong local focus on regionally confined containment measures may not be helpful. Our findings suggest that for effective disease mitigation similar rules should be implemented in neighboring regions. If a regional hotspot of infections emerges, it would thus make sense to follow the same disease mitigating strategy both in the hotspot and its neighboring regions or alternatively make sure that no in- and out-mobility from the hotspot takes place.

Some caveats in interpreting the impacts of the policy actions as causal effects should be highlighted, though. First, it could be conceivable that regionally varying test capacities influence the number of tested people and hereby reported infection cases. However, in view of homogenous rules of testing across German regions, this issue might play a more important role for international comparisons (cf. Mitze et al. 2020). Second, our estimates are conditional on the sequence of individual measures. Because of a lack of observational facts, we refrain from speculating about potential effects from other sequences. In this context we would also like to stress that we are only able to identify the overall effect of a public health measure, but not its underlying mechanisms. Specifically, sudden and unprecedented measures such as school closures or strict contact restrictions may lead to strong behavioural adjustments that increase observed effects of a particular measure, specifically if it comes first in the sequence of events (e.g. with the closing of schools the local population already adjusts is shopping behaviour even if shops are still open etc.). This type of behaviour spillover is also referred to as Hawthorne effect.

Third, we do not explicitly cover all implemented containment measures in our analysis, particularly if these took place on a voluntary basis such as home office work for many white-collar workers in Germany. In the latter case, it is very difficult to construct a meaningful treatment dummy with a precisely defined starting date at the regional level. Moreover, many firms allowed employees to switch to home office work already in late February and early March. We thus argue that the gradual increase of home office work during our sample period is not expected to confound our treatment effect estimates for measures taken from mid-March onwards. Further, growth in the utilization of home office work throughout the sample period should be captured by the region-specific time trends in our spatial DiD approach. Another measure that is not explicitly covered here are international travel restrictions (or more specifically: international travel warnings). Since these restrictions are typically issued by the German Foreign Office and are equally binding for all German districts, we argue that effects resulting from changes in international travel warnings should be fully captured by the included time-fixed effects.

Finally, reverse causation is always a concern when policymakers just react to accelerating infection rates. We believe that the design of our study is suitable to avoid a noticeable endogeneity bias. However, this strictly applies only to the measures being in the specific chronological order of their implementation (as outlined above). Thus, the containment effect of closure of schools and daycare facilities is backed under the condition that the intervention takes place before contact restrictions and the mask duty are introduced. The same holds for the unstable effect of restaurant closure. In this context, particularly the estimated face mask effect appears to be significant and well grounded. Also, the continued imposition of the contact restrictions by the states turns out to be well justified. In particular, a low impact of this measure on infection growth would question the efficacy of such severe constraints. Considering the strong treatment effects of contact restrictions and mandatory face mask wearing, the interaction between both measures should become a future research issue (Seres et al. 2020).

An optimal instrument mix has to take account of the economic costs of tightening or easing public health actions to curb the spread of the coronavirus. While detailed cost-benefit analyses are scarcely available yet, some studies aim to tackle this issue for selected measures. Fuchs-Schündeln, Kuhn and Tertilt (2020) find strong labor market effects in the wake of school and kindergarten closure. They estimate that 8.4 percent of total working hours will be lost, corresponding to 11.7 million employed persons in short-time-work. Beyond this short-term effect, intergenerational mobility and gender equality in the workplace is likely negatively affected. The macroeconomic impact of this public health measure is considered large. Based on simulation, Bürgi and Gorgulu (2020) assess a 2 percentage points larger drop in Q2 GDP in the United States for social distancing at the level of most European countries.

Substantial GDP impacts of closures of retail stores, restaurants and other business outlets are found by Basurto et al. (2020) under various degrees of easing restrictions. Because the lack of robust treatment effects of these measures, their efficiency must be strongly challenged. Economic effects of several interventions are examined by Stannard, Steven and McDonald (2020) for the economy of New Zealand. While a moderate decline in DGP of 0.7 percent is estimated for the ban of mass gatherings and closure of public venues, restricting domestic travel may induce a large reduction up to 6 percent. Economic effects of social-distancing policies are investigated by Strong and Welburn (2020) and Fernandéz-Villaverde and Jones (2020). Compared with other common measures, the implied economic costs for community masks seem comparatively low. However, further cost-benefit analyses are needed to verify whether wearing face masks is not only an effective but also cost-efficient measure for fighting Covid-19. Similar considerations need also be done for other measure taken in the past (and future) to fight the Covid-19 pandemic.

## Data Availability

Data is public

## Appendix A: Implementation dates of containment measures

## Appendix B: Day of Diagnosis and Day of Reporting

German authorities (RKI, 2020a) measure the number of reported infections by day of reporting and by day of first appearance of symptoms. Both ways of reporting have their advantages and disadvantages. Data by date of reporting is cyclical over the week. Incidences systematically fall over the weekend and are highest around the mid of the week. At weekends, testing is reduced and local health authorities are usually closed (RKI, 2020b). As reporting does not relate one-to-one to the spread of infections, employing reporting data introduces additional measurement uncertainty. This would suggest working with incidence data by day of diagnosis.

However, data by day of diagnosis has disadvantages as well. Not all cases reported to the RKI come with the date of diagnosis. In many cases, we only know the day of reporting. As the figures show, more than one fourth of the data of the data is still by day of reporting. For these cases, knowledge on the day of first appearance of symptoms is missing. The share the data provided by day of reporting seems to increase over time.

There is one great advantage of using data by day of diagnosis. The median delay between infection and appearance of symptoms is 5.2 days with 95% lying between 2 and 12 days (Linton et al., 2020 and Lauer et al., 2020). When we are interested in the effect of some containment measure on the number of infections and use data by day of diagnosis, we would expect that the about 50% of the effect is visible five days after the measure was implemented. When we use data by day of reporting, we need an additional delay of 5 to 6 days for the patient to visit a doctor and the reporting procedure to reach this threshold. This enters further imprecision in measurement.

We therefore need to choose between a dataset that may be more precise but is not homogenous (day of first symptoms) and a dataset where we need to add additional days for reporting. In order to employ the comprehensive RKI dataset, we relate infection cases to the day of reporting. With a delay of 7 days between infection and reporting, about 25% of infection cases are expected to be statistically visible. As in accompanying work (Kleyer et al. 2020) we capture cyclicality over the week by the day of the week effect.

## Appendix C: Day of the Week Effect

Figure 8 exhibits the pattern of the day of the week effect with Sunday as the reference day. Particularly because of limited testing on weekends, figures begin to rise at the start of the week. The highest growth of incidence rates of additional 7 ½ percentage points is observed on Wednesday. Growth rates are almost as high on Tuesday and Thursday. The low weekend effect is also clearly visible for Saturday. All weekday impacts are highly significant.

## Footnotes

2 Early voluntary cancellations of mass events pose an additional problem in modeling measure-specific contaiment effects.

3 We use a distance-based weights matrix in robustness analysis (sect. 6).

4 Development trends of infection cases vary considerably across German (cf. Donsimoni et al. 2020).

5 Daily seasonalities occur as troughs at the weekend and peaks in the middle of the weak in infection cases (RKI 2020b).

6 Delgado and Florax (2015) introduce a spillover parameter ρ that implies θ_k_ = ρ·γ_k_. This relation can be helpful for estimating the combined effect in case of multicollinearity.

7 When the common factor assumption θ_k_ = -λ·β_k_ holds for all k, endogenous spatial spillovers can be captured by the SEM model (cf. LeSage and Pace 2009, 164-165).

8 The GNS model is also termed Manski model (Floch Le Saout 2018, 169). We use the GNS model (Manski model) for robustness checking.

9 Only in exceptional cases an early adoption of a measure took place in single regions. In these exceptions, an additional control is given by the inclusion of region-specific time trends (Kolak and Anselin 2020).

10 As the 25% percentile of total delay coincides with the selected lag, infection effects are expected to be visible in the registered cases after one week (Appendix E, Mitze et al. 2020, Appendix E). In sect. 6 we examine the sensitivity of treatment effects with respect to alternative lag selections.

11 If we assume that contact restrictions were also effective in Rhineland-Palatinate and Thuringa the day following their announcement, estimated reponse coefficient of this policy action slightly drops to 0.12 in absolute terms. Thus, a stronger impact of this measure on growth rates of infections is associated with its actual implementation.

12 The SAR model only very indirectly captures policy interventions in neighbouring regions running through changes in the spatial lag of the endogenous variable as a “catch-all” term.

13 Mitze et al. 2020 find that with total delays between 5 and 10 days roughly 5 to 50% of infections are visible in RKI data.

14 A different identification approach that analyses the role of schools as drivers of the pandemic development in Germany is given in von Bismarck-Osten, Borusyak and Schönberg (2020). The authors mainly assess the infection effects related to school closure and opening by exploiting spatio-temporal differences in the start and end dates of school summer (and autumn) holidays across federal states. The empirical findings do not indicate a significant contribution of regular school closures on Covid-19 incident rates. However, while this finding is an important complementary statement to our approach, it can be argued that behavioural adjustments in response to regular, i.e. holidays, and sudden school closure, as in our case, are very different from each other and may thus affect regional infection dynamics in a very heterogeneous way. On top, intervention effects may alter along a region’s position on the pandemic curve. Effects can be expected to be larger during a specific wave (as in spring 2020) compared to a situation with very low to moderate overall infection levels (as in summer 2020).

